# Mathematical modeling of mpox: a scoping review

**DOI:** 10.1101/2022.12.28.22284007

**Authors:** Jeta Molla, Idriss Sekkak, Ariel Mundo Ortiz, Iain Moyles, Bouchra Nasri

## Abstract

**Background:** Mpox (monkeypox), a disease historically endemic to Africa, has seen its largest outbreak in 2022 by spreading to many regions of the world and has become a public health threat. Informed policies aimed at controlling and managing the spread of this disease necessitate the use of adequate mathematical modelling strategies.

**Objective:** In this scoping review, we sought to identify the mathematical models that have been used to study mpox transmission in the literature in order to determine what are the model classes most frequently used, their assumptions, and the modelling gaps that need to be addressed in the context of the epidemiological characteristics of the ongoing mpox outbreak.

**Methods:** This study employed the methodology of the PRISMA guidelines for scoping reviews to identify the mathematical models available to study mpox transmission dynamics. Three databases (PubMed, Web of Science and MathSciNet) were systematically searched to identify relevant studies.

**Results:** A total of 5827 papers were screened from the database queries. After screening, 35 studies that met the inclusion criteria were analyzed, and 19 were finally included in the scoping review. Our results show that compartmental, branching process, Monte Carlo (stochastic), agent-based, and network models have been used to study mpox transmission dynamics between humans as well as between humans and animals. Furthermore, compartmental and branching models have been the most commonly used classes.

**Conclusions:** There is a need to develop modelling strategies for mpox transmission that take into account the conditions of the current outbreak, which has been largely driven by human-to-human transmission in urban settings. In the current scenario, the assumptions and parameters used by most of the studies included in this review (which are largely based on a limited number of studies carried in Africa in the early 80s) may not be applicable, and therefore, can complicate any public health policies that are derived from their estimates. The current mpox outbreak is also an example of how more research into neglected zoonoses is needed in an era where new and re-emerging diseases have become global public health threats.

## Background

### Rationale

Zoonotic diseases continue to be major public health threats around the world, as exemplified by a global outbreak of monkeypox (the authors note that, following the World Health Organization’s recommendation of new name, this study will hencefort refer to the disease as”mpox” [1]) that began in early May of 2022 when a mpox case not linked to travel to endemic countries, was first reported in the UK [19]. Mpox is a disease caused by the double-stranded DNA virus *monkeypox* from the *Poxviridae* family). As of October of 2022, this mpox outbreak has accounted for more than 63,000 cases in 106 different locations [7], while being declared a”public health emergency of international concern” by the World Health Organization [29] due to its extensive geographical distribution and its occurrence at a time when healthcare systems around the world continue to experience stress due to the ongoing COVID-19 pandemic [23].

Although mpox is considered a mild and self-limiting disease, it is known that it can be severe in the case of children, pregnant women, and immunocompromised individuals [22, 30]. In the past,the majority of previous mpox outbreaks were localized in neglected communities of West and Central Africa, where the disease is endemic [20, 28, 33].

Due to global vaccination efforts against smallpox between 1967 and 1979, population immunity to mpox existed as the smallpox vaccine offered a certain degree of protection. However, vaccination ceased with the eradication of smallpox in 1979 and the immunity against mpox of those vaccinated is now waning [34, 24]. The combination of epidemiological evolution of the virus and loss of vaccine protection is cause for concern in the ongoing global mpox outbreak.

Public health policies concerned with planning and response to the mpox outbreak require the use of adequate disease modelling strategies. This fact has been continually demonstrated during the ongoing COVID-19 pandemic where mathematical models of disease transmission have played a critical role to inform public health policies [35]. On the other hand, mpox has historically been a neglected zoonotic disease and consequently, mathematical models of mpox transmission are limited in volume and application.

### Research Questions

The objectives of this scoping literature review are: 1) to generate a repository of models and parameters/data that have been used to understand the dynamics of mpox in order to address current epidemiological questions, and 2) identify the limitations of available modelling strategies in the context of the current mpox outbreak.

## Methods

This scoping review followed the Preferred Reporting Items for Systematic reviews and Meta-Analyses (PRISMA) guidelines and checklist [25]. The checklist is provided in Figure 1. With the exception of the “critical appraisal of individual sources” (since the main goal is to provide and classify a repository of models and parameters/data) all items in this checklist were included in this study. A detailed description of each item is provided below.

**Figure 1:**
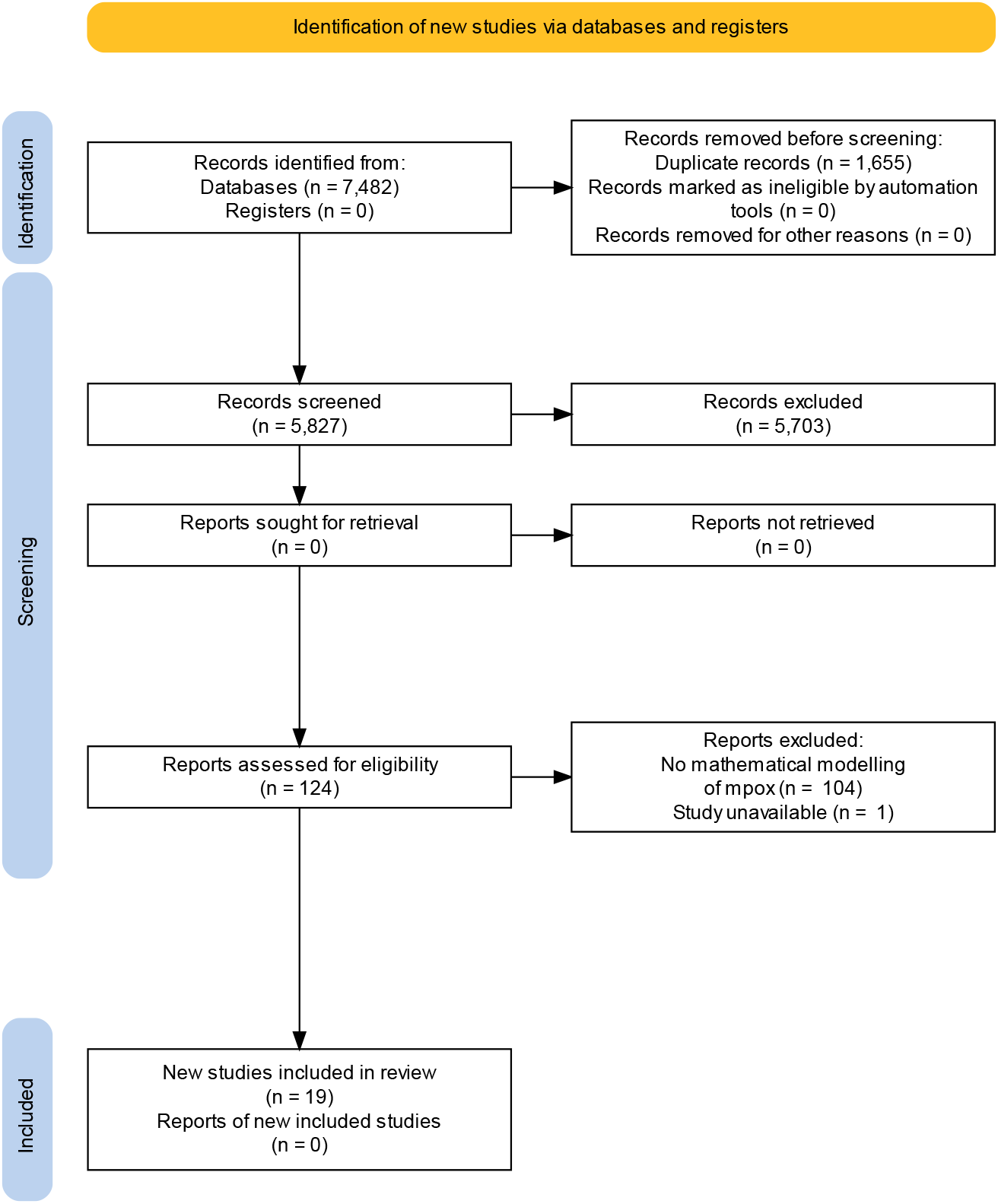
Diagram of Identification of new studies.

### Eligibility Criteria

Inclusion criteria were original research peer-reviewed articles in English with publication date before October 19, 2022 that used a mathematical modelling approach to describe the transmission dynamics of the mpox virus, its genus (*Orthopoxvirus*) or family (*Poxvirus*), and that focused on disease spread among humans, animals, or between animals and humans. No geographical restrictions were considered.

### Information Sources

Three databases were searched: PubMed, Web of Science, and MathSciNet. PubMed was chosen as it contains references and abstracts on subjects related to the life sciences and biomedicine which were areas of major interest in the context of this study. To complement PubMed results, Web of Science was searched to broaden the scope beyond life sciences and biomedicine. Finally, MathSciNet was chosen because of its focus on cataloguing papers in mathematical sciences, thereby identifying modelling literature not found in the other two databases.

### Search Strategy

The searches were conducted on October 19, 2022. The search query contained all combinations of {`keyword1`} AND {`keyword2`} where the set of `keyword1` included *monkeypox, Orthopoxvirus, Poxvirus, Poxviridae, MPV, MPXV*, and *hMPXV*, while the set of `keyword2` included *model, simulation, computation, travelling wave, machine learning, equation, process*, and *math*. The specific search method using these keywords for each database can be found in the Appendix. The decision process for eligibility was conducted by four of the authors in collaboration (JM, IS, IM, BN).

### Selection of Sources of Evidence

Four of the authors (JM, IS, IM, BN) were in charge of the first round of the screening process. Results of the search in each database were imported into the systematic review software *Covidence* [14], where duplicates were removed and the screening process took place. In the first round of screening, authors independently reviewed the title and abstract of each recovered paper and assigned a ‘yes’, ‘no’, or ‘maybe’ vote for the inclusion of the study in the next screening phase. A paper was accepted to the second phase if it received two ‘yes’ votes, and rejected if it received two ‘no’ votes.

Papers voted ‘maybe’ or that had non-unanimous decisions were evaluated by a third author and a final decision was made. Exclusions at this round of screening were based on the assessment if a paper fit the criteria of mathematical modelling. For example, literature on infectious diseases where animals are used as a proxy for humans refers to the studied animal species as a ‘model’. Such definition of ‘model’ is distinct from mathematical definition of ‘model’ (i.e., description of a system using mathematical concepts) and such studies were therefore excluded.

All authors contributed in the second round of screening, where least two of them read the full-text of each study. Each author assigned an ‘include’ or ‘exclude’ vote. A study was included for analysis if it received two ‘include’ votes, and it was excluded from further analysis if it received two ‘exclude’ votes. An ‘exclude’ vote was justified with one of the following two exclusion criteria: *no mathematical modelling of mpox* (which encompassed studies focused on animal models, genome studies, statistical analyses, models for other diseases, and studies where no model was presented) and *study unavailable* (when the link provided did not redirect to the corresponding study).

### Data Charting Process and Data Items

A narrative review was given for each of the included studies in a spreadsheet with the following information collected: electronic link to publication, year of publication, geographical region, type of model and data used, parameters of the model, as well as detailed notes about methods and results. Additional comments (e.g., limitations of the study) from the reviewers were also included.

### Synthesis of Results

The studies were organized according to the class of mathematical model presented in each case using three categories. The first category corresponded to models where a compartmental modeling framework was used to describe the transmission dynamics between human and/or animal populations. The second category corresponded to branching process models, while the third category included studies that used a modeling approach that did not fit in any of the previous two categories.

## Results

### Selection of sources of evidence

The database queries resulted in 7482 papers meeting the selected criteria. Following the removal of duplicates, 5827 studies were moved to the screening phase. The first stage of screening resulted in 124 studies being retained, and the second stage of screening resulted in 35 studies for which the full text was analyzed. Following full-text analysis, 19 studies were included in this scoping review. A PRISMA diagram summarising the stages of the screening process is presented in Fig. 1.

### Characteristics of Included studies

In Table 1 the included studies are sorted chronologically by publication date while including the region of study (geographical area from which data was obtained to estimate parameters or to validate the model) and the type of model used in each case. According to the search query parameters, the earliest relevant paper on mathematical modelling of mpox transmission was published in 1987 and the latest in 2022. In seven of studies the geographical regions of interest were located in Africa, from either the Democratic Republic of the Congo (previously Republic of Zaire), Central and West Africa (the Congo Basin is located in Central Africa), and Nigeria. This result is consistent with the endemic spread of the virus in these regions prior to 2022 [33]. Three studies reported regions of interest in Europe (Belgium, Austria, and the United Kingdom), and nine studies reported no geographical region of interest.

**Table 1:** Date of publication, region of study, and type of model considered in the included papers.

| Date of publication | Region of study | Type of model |
| --- | --- | --- |
| 1987 | Democratic Republic of the Congo (previously Republic of Zaire) | Monte Carlo stochastic model [16] |
| 1999 | Democratic Republic of the Congo (previously Republic of Zaire) | Branching process [11] |
| 2003 | No geographic region identified | Branching process [2] |
| 2012 | No geographic region identified | Compartmental [4] |
| 2012 | No geographic region identified | Compartmental [38] |
| 2013 | Democratic Republic of the Congo (previously Republic of Zaire) | Branching process [36] |
| 2014 | Democratic Republic of the Congo (previously Republic of Zaire) | Branching process [5] |
| 2015 | Central Africa | Branching process [21] |
| 2020 | Central and West Africa | Compartmental [3] |
| 2020 | No geographic region identified | Agent-based [6] |
| 2020 | No geographic region identified | Branching process [27] |
| 2021 | No geographic region identified | Compartmental [31] |
| 2022 | Nigeria | Compartmental (fractional order) [32] |
| 2022 | Belgium | Network model [8] |
| 2022 | No geographic region identified | Compartmental [39] |
| 2022 | Austria | Compartmental [37] |
| 2022 | No geographic region identified | Compartmental (stochastic) [18] |
| 2022 | United Kingdom | Branching process [10] |
| 2022 | No geographic region identified | Compartmental (fractional order) [9] |

### Results of individual sources of evidence

The analysis found that compartmental and branching process models are the main modelling framework used to study the transmission dynamics of mpox, with nine studies using the first model class and seven studies using the latter. Additionally, Monte Carlo (stochastic), agent-based, and network models have also been used, albeit with relatively less frequency. Next, a summary of the major findings of the studies within each modelling framework is presented.

#### Compartmental Models

Of the 19 studies retained after the second screening stage, nine used a compartmental model to describe the transmission dynamics of the mpox virus in a population consisting of humans and non-humans (Table 1). These nine studies analyzed the transmission dynamics of mpox between humans, as well as between humans and animals using the standard susceptible-infected-recovered (SIR) classes [17]. However, although most studies considered an “exposed” compartment to model the spread of the disease, in some cases the authors added a “quarantined” or “isolated infected” compartment to study the effect of control measures [31, 18, 32]. Other compartments that were added in some studies to account for various scenarios of disease spread include compartments for those vaccinated [38, 37], those clinically ill [32], and compartments to model different stages of mpox where infection can occur (prodromal and rash phases) [39]. Two of the nine compartmental studies considered an extension of the classical model by introducing fractional order time-derivative [9, 31] and one considered a stochastic extension with Lévy noise [18]. Furthermore, the non-human population in the compartmental studies included monkeys and/or rodents [4, 31, 32], squirrels [3], or all of the species that can carry mpox [38].

The compartments considered in each of the compartmental model studies can be found in Table 2, whereas the values of selected parameters reported in the same studies can be found in Table 3. For generality, Table 3 omits parameters that are geographically-dependent (such as birth rate) and parameters that are related to transmission solely within animal populations; researchers interested in such parameters can refer to the original studies directly.

**Table 2:** Compartments used in the compartmental models for mpox.

| Compartments | Definition |
| --- | --- |
| $S_S, E_S, I_S, R_S, S_h, E_h, I_h, V_h, R_h$ | Susceptible ($S$), exposed ($E$), infected ($I$), vaccinated ($V$), and recovered ($R$). The subscripts $S$ and $h$ stand for compartments in the squirrel and human population, respectively [3]. |
| $S_r, I_r, R_r, S, I_m, R_m, I_h, A_h, I_{hm}, A_{hm}, R_h, R_{hm}, R_{am}$ | Susceptible rodents ($S_r$), infected rodents ($I_r$), recovered rodents ($R_r$). Susceptible $S$ humans, infected with mpox ($I_m$), infected with HIV ($I_h$), HIV infected and in the AIDS stage of disease progression ($A_h$), infected with HIV and mpox ($I_{hm}$), dually infected with mpox and HIV and in the AIDS stage of disease progression ($A_{hm}$), recovered from mpox ($R_m$), recovered HIV and mpox ($R_{hm}$), recovered from mpox and are infected with HIV in the AIDS stage of disease progression ($R_{am}$) [4]. |
| $S_r, E_r, I_r, S_h, E_h, I_h, Q_h, R_h$ | Susceptible ($S$), exposed ($E$), infected ($I$), quarantined ($Q$), and recovered ($R$). The subscripts $r$ and $h$ describe the compartments in the rodent and human population, respectively [31]. |
| $S_M, I_M, R_M, S_{hi}, I_{hi}, V_{hi}, R_{hi}$ | Susceptible ($S$), infected ($I$), vaccinated ($V$), and recovered ($R$). The subscripts $r$ and $h$ account for the compartments in the animal and human population, respectively. The $i$ population describes whether an individual belongs to the juvenile or adult population [38]. |
| $S'_h, E'_h, I'_h, C'_h, R'_h, S'_r, E'_r, I'_r$ | Susceptible ($S'$), exposed ($E'$), infectious ($I'$), clinically ill ($C'$), recovered ($R'$). Subindices indicate humans ($h$) or rodents ($r$) [32]. |
| $S_{h1}, E_{h1}, I_{h1}, P_{h1}, Q_{s1}, S_{h2}, E_{h2}, I_{h2}, P_{h2}, Q_{s2}, R_h, Q_h, S_r, E_r, I_r, R_r$ | Susceptible ($S$), exposed ($E$), infected at prodromal phase ($P$), infected at rash phase ($I$), isolated and infectious ($Q_h$), isolated and susceptible ($Q_s$), recovered ($R$). Human sub-populations are modeled as low-risk ($h1$), and high-risk ($h2$), rodents are indicated by subindex $r$ [39]. |
| $S_{A,t}, S_t, I, R$ | Proportion of individuals within the population at risk at time $t$ that were born on year $A$ at risk of infection considering smallpox vaccination status ($S_{A,t}$), total proportion of susceptible individuals ($S_t$), infected ($I$), recovered ($R$) [37]. |
| $C_{1,h}, C_{2,h}, C_{3,h}, C_{4,h}, C_{1,m}, C_{2,m}$ | Sensitive ($C_1$), infected ($C_2$), isolated infected ($C_3$), recovered ($C_4$). Subindices indicate humans ($h$) or animals ($m$) [18]. |
| $S_h(t), E_h(t), I_h(t), Q_h(t), R_h(t), S_r(t), E_r(t), I_r(t)$ | Susceptible ($S(t)$), exposed ($E(t)$), infectious ($I(t)$), recovered ($R(t)$). Subindices indicate humans ($h$) or rodents ($r$) [9]. |

**Table 3:**
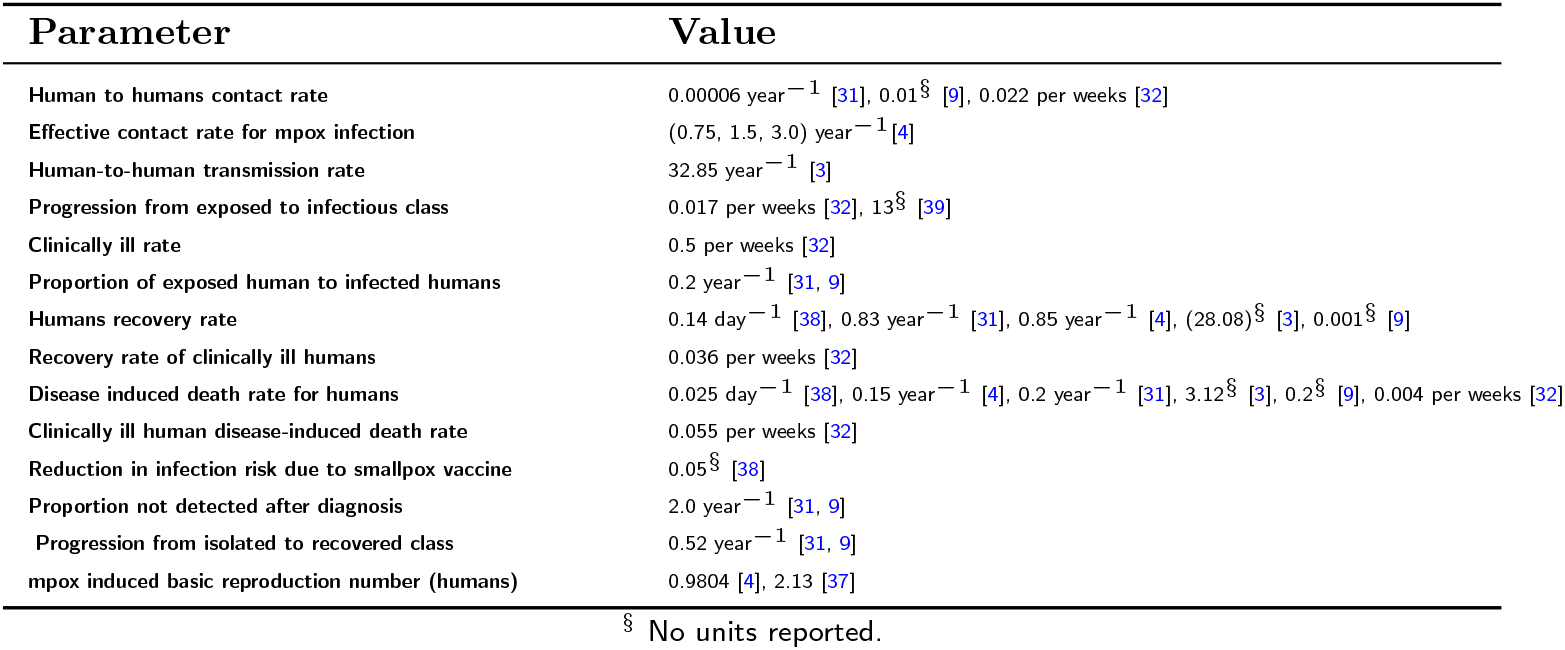
Selected parameters used in the compartmental models for mpox.

| Parameter | Value |
| --- | --- |
| Human to humans contact rate | $0.00006 \text{ year}^{-1}$ [31], $0.01^{\S}$ [9], $0.022 \text{ per weeks}$ [32] |
| Effective contact rate for mpox infection | $(0.75, 1.5, 3.0) \text{ year}^{-1}$ [4] |
| Human-to-human transmission rate | $32.85 \text{ year}^{-1}$ [3] |
| Progression from exposed to infectious class | $0.017 \text{ per weeks}$ [32], $13^{\S}$ [39] |
| Clinically ill rate | $0.5 \text{ per weeks}$ [32] |
| Proportion of exposed human to infected humans | $0.2 \text{ year}^{-1}$ [31, 9] |
| Humans recovery rate | $0.14 \text{ day}^{-1}$ [38], $0.83 \text{ year}^{-1}$ [31], $0.85 \text{ year}^{-1}$ [4], $(28.08)^{\S}$ [3], $0.001^{\S}$ [9] |
| Recovery rate of clinically ill humans | $0.036 \text{ per weeks}$ [32] |
| Disease induced death rate for humans | $0.025 \text{ day}^{-1}$ [38], $0.15 \text{ year}^{-1}$ [4], $0.2 \text{ year}^{-1}$ [31], $3.12^{\S}$ [3], $0.2^{\S}$ [9], $0.004 \text{ per weeks}$ [32] |
| Clinically ill human disease-induced death rate | $0.055 \text{ per weeks}$ [32] |
| Reduction in infection risk due to smallpox vaccine | $0.05^{\S}$ [38] |
| Proportion not detected after diagnosis | $2.0 \text{ year}^{-1}$ [31, 9] |
| Progression from isolated to recovered class | $0.52 \text{ year}^{-1}$ [31, 9] |
| mpox induced basic reproduction number (humans) | $0.9804$ [4], $2.13$ [37] |

<sup>§</sup> No units reported.

The full-text review process of the compartmental studies resulted in grouping the main findings of the papers in two categories: 1) the effect of mitigation measures and 2) stability analysis of equilibrium points.

##### Effect of mitigation measures

Five of the nine analyzed compartmental studies investigated the impact of pharmaceutical and non-pharmaceutical interventions on mitigating the spread of mpox. Tchuenche and Bauch [38] numerically studied the effect of culling an animal host and their findings suggest that the impact of culling strongly depends on the details of demography and epidemiology in the animal reservoirs that sustain it. For some parameter values, increased culling could actually have the counterproductive outcome of increasing mpox infections. Peter et al. [31] showed that isolating infected individuals is crucial for controlling the spread of mpox, while Yuan et al. [39] studied the effect of isolation and contact tracing and concluded that contact tracing was more important in the high risk group to contain the spread of the disease. The authors in [3] and [37] focused on vaccination as a measure to control the spread of the disease. Bankuru et al. [3] showed that if mpox is in a semi-endemic equilibrium, then it can be controlled and eradicated using vaccination, while Spath et al. [37] concluded that partial immunization would be insufficient to contain a mpox outbreak in the absence of other control measures.

##### Stability analysis of equilibrium points

Six of the nine compartmental model studies investigated the stability of the mpox-free and endemic equilibrium points (i.e., the conditions under which the disease persists or dies out). This was done using a classical SIR framework (cf. [4, 31, 3]) as well as a fractional compartmental framework (cf. [32, 9]). Both approaches identify a reproduction number in their model and derive the conditions under which the mpox disease is eradicated (reproduction number is smaller than 1) and when it persists (reproduction number is larger than 1). Stability was also investigated by Khan *et al*., using a stochastic compartmental model in a mean-field limit [18]. They concluded that the conditions for eradication or persistence are distinct in contrast to deterministic models where a single quantity, the reproduction number, determines the disease trajectory.

#### Branching Process Models

Branching process models were used in seven of the 19 studies analyzed. Six of these studies employed offspring distributions to model the spread of the disease [11, 5, 36, 2, 21, 10], and five studies used data from empirical epidemiological studies to assess the accuracy of their models [11, 5, 36, 21, 10]. Interestingly, three studies used the same dataset from mpox transmission in Zaire in the early 1980s to validate the results of their models [11, 5, 36], although they assumed different offspring distributions. The offspring distributions, details about the geographical location, time span, and source of the empirical data used to determine the accuracy of the models are presented in Table 4.

**Table 4:**
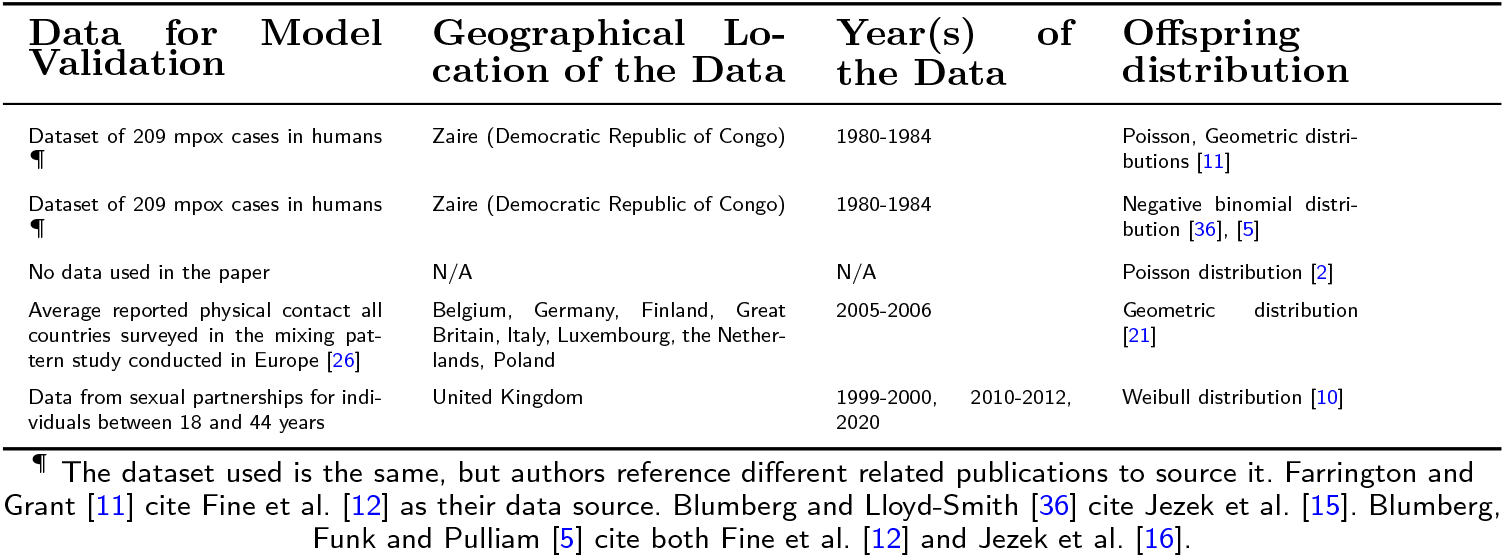
Offspring distributions used in branching process models for mpox

Based on the full-text review of the branching process studies, the studies were divided in two groups to analyze their major findings: 1) studies that obtained estimates of reproduction and transmission values (basic reproduction number *R*_0_, effective reproduction number *R*_eff_, heterogeneity in infectionusness *k*) in different scenarios, and 2) studies that focused on model development to address specific questions of disease spread. The results of the studies within each group are presented next.

##### Estimates of reproduction and transmission

Four studies focused on obtaining estimates of reproduction and transmission considering different scenarios. Kucharski et al. [21] estimated *R*_0_ considering age stratification, finding that individuals above 20 years of age had reduced susceptibility of infection by a factor of 0.4. Blumberg and Lloyd-Smith [36] obtained estimates of *R*_0_ and *k* using only the total number of infected cases (chain size) showing that there was good agreement between the values obtained with this method and those obtained using contact tracing data. On a follow up study, Blumberg et al. analyzed if different levels of transmission existed between primary cases (animals to humans) and secondary cases (among humans) by calculating *R*_eff_ and *k* in each case [5], demonstrating that the levels of transmission between primary and secondary cases were not significantly different. Finally, Endo et al. [10] calculated *R*_0_ for mpox transmission between men who have sex with men (a group that has seen a high number of cases in the current outbreak), showing its value was substantially above 1 in this demographic group. Importantly, the first three studies assumed that mpox was a a “sub-critical disease”, effectively meaning that its spread was limited by *R*_0_ *<* 1. Only the last study accounted for conditions present in the current outbreak. The reported values of *R*_0_, *R*_eff_, and *k* in these studies are presented in Table 5.

**Table 5:** Reproduction and transmission numbers reported for branching process models.

| Parameter | Value |
| --- | --- |
| Basic reproduction number ($R_0$) | 0.3(0.21 – 0.42)¶ [36], $10^2$ [10] |
| Effective reproduction number ($R_{\text{eff}}$) | 0.08(0.02 – 0.22)¶ [21], 0.3, [5] |
| Heterogeneity in infectiousness ($k$) | 0.36(0.14 – 2.57)¶ [36], 0.4 [5] |

¶ 95% confidence interval reported for the value.

##### Studies of model development

The remaining three branching process studies focused on developing models that could help estimate disease spread when different factors were considered. Farrington et al. aimed to estimate the number of generations to extinction for mpox (considered as a sub-critical branching process) by modelling the spread of the disease as Poisson or geometric distributions in the presence of partial vaccination and finding that both offsprings distributions resulted in close approximations to empirical data [11]. Antia et al. developed a model to analyze how a virus with an initial value of *R*_0_ below 1 can be affected by mutations and ecological factors during the chain of transmission such that at some point the value of *R*_0_ approaches 1, thereby causing an epidemic [2]. The authors found that the probability of infection with an evolved strain was highly sensitive to the value of *R*_0_, and is approximately linearly dependent on the mutation rate. Finally, Mummah et al. developed a model to study how disease control policies (such as isolation) affect the value of the reproduction number, and thereby transmission, and also provided guidelines on control policy implementation considering costs and limited resources [27].

#### Other Models

The remaining three studies in Table 1 used models that were not compartmental or branching process to analyze mpox transmission. Jezek et al. [16] built a Monte Carlo stochastic model to simulate the chain of mpox transmission from patients to their exposed close contacts while keeping track of the generation order for successive secondary cases [16]. The model showed an increase in new secondary cases in the absence of smallpox vaccination, but without reaching pandemic proportions (the study used the same dataset from Zaire from early 1980s).

Brainard and Hunter [6] designed an agent-based model (ABM) to account for disease spread of three diseases, including mpox. In this study, the agents represented a certain percent of independent individuals in a population that were infected with mpox and that shared health advice with other agents (either useful or harmful). The authors found that restricting harmful advice consequently mitigated its effect in the disease outcomes.

Finally, Van Dijck et al. [8], used a network model to describe the transmission of mpox among men who have sex with men using behavioral data from Belgium. This study found that a proportion of undetected cases is 50% and results in an 8-fold increase in the actual number of cases compared to the estimate obtained from the diagnosed cases.

## Discussion

Different modelling frameworks (compartmental, branching process, Monte Carlo stochastic, ABM, network) have been used to study mpox transmission. It is important to discuss the applicability of these modelling frameworks in the context of the current outbreak by considering their limitations, and identifying the gaps that need to be addressed by new models.

The SIR model was employed in all the compartmental model studies (Table 2). The implementation fo this model is straightforward as it requires a small number of parameters, which makes it a convenient framework to study the mpox transmission dynamics and evaluate the effectiveness of various interventions. However, a limitation of this model is that it assumes homogeneous mixing of the population (i.e., all individuals in the population are assumed to have an equal probability of coming in contact with one another). Although, three of the nine compartmental model studies divided the population in different age groups [38, 37], and risk groups [39], and assumed different contact probabilities among these groups, none of the nine studies included spatial structure or took into account that individuals in the population can be infected only by a constrained set of other individuals since the majority of contact occurs within limited networks. Therefore, it is important that future studies consider spatial or social structures in order to contextualize the social and geographical conditions of the current mpox outbreak. The target populations and the contact structure of the community also pose challenges for SIR modelling, making necessary in future studies the use of models that adequately consider the heterogeneity in transmission [13].

Another limitation of the compartmental models analyzed in this study pertains to the data sources used to estimate the model parameters (Table 3), where in some cases parameters had a wide range of variation. Authors often reported values to be either assumed or estimated using available literature. However, in some situations the exact rationale used to derive the values was not clear, and additionally, there was a lack of important information to assess validity (such as units). In reality, it is quite possible that the magnitude of the parameters is different from the numbers reported due to the uncertainty associated with their estimation. This could be partly attributed to assumptions in the literature, but it is also due to mpox’s history as a neglected and understudied disease. Therefore, it is critical that more accurate parameters are estimated using empirical studies of mpox epidemiology in human populations. Accurate parameters are required to facilitate the development of more sophisticated compartmental models that are applicable to the current mpox outbreak.

In the case of the branching process models, a major limitation in the applicability of the models and estimates from the studies in the context of the current outbreak pertains to the assumption of mpox as a self-limiting disease (*R*_0_ *<* 1). It is clear that in the current outbreak *R*_0_ has exceeded the self-limiting threshold because the spread continues at the human level, and therefore the assumptions made in a self-limiting scenario are no longer applicable.

Such limitation can also be identified in the data used in some of the analyzed studies, which used the same dataset of mpox transmission in Zaire (Democratic Republic of Congo) from the early 1980s to validate the models (Table 4). This dataset reported 209 cases of mpox transmission collected over 5 years in a country that, in 1982, had a population of around 8.4 million [16]. Therefore, the rates of infection, level of transmission, and the overall assumptions made in studies that used this data to estimate the values of *R*_0_ and *R*_eff_ are likely not applicable to the current outbreak [27].

The usage of data collected in the 1980s in literature published 30 years later is yet another indication of how mpox has historically been a neglected zoonosis, pointing to the need of current field studies that allow for the adequate estimation of *R*_0_ and *R*_eff_. However, it worth mentioning that although the most recent branching process study did use recent data to fit the model [10], the type of data used (sexual contacts in the United Kingdom) makes the applicability of the results of the study difficult within a broad population context.

Some of the branching process studies focused on model development lacked reproducibility (i.e., no data used to validate the model assumptions), such as in Antia [2], and Mummah et al. [27]. In both cases, the authors provided only a purely theoretical approach and very succint details on the assumptions on the model, which greatly complicates any attempt to validate the model assumptions with data that could be obtained for the current outbreak.

However, a valuable contribution from recent branching process modelling strategies is the use of contact pattern data collected from a large geographical area [21]. Such a methodology could be valuable in assessing mpox spread over a large geographical area contrasting the survey and payment-based methodology used in the past, which would be economically prohibitive in the case of the current outbreak. Three of the studies in Table 1 did not use compartmental or branching process models to analyze mpox transmission. The Monte Carlo stochastic model of Jezek et al. [16] presented an interesting approach that did not follow the deterministic nature of compartmental or branching process models. This could be useful for modelling random noise due to geography or other heterogeneities within the curent outbreak. However, the effective implementation of a stochastic model would require comprehensive transmission data and many stochastic realizations. The study from Jezek et al. used the Zaire dataset from the 1980s and computed results based on simulated series repeated only 100 times.

The ABM model approach of Brainard and Hunter [6] could be advantageous to study mpox transmission in the current outbreak as it can assist model developers and users in managing adaptation, spatial structure, and heterogeneity (three specific complex difficulties for researchers and decision-makers). However, this class of models are known to be time and computationally-intensive due to the necessity of tracking all of the agents in the model. For example, the model used by Brainard and Hunter used 1600 agents for the current mpox outbreak which is significantly less than the susceptible populations of most geographical areas. It is possible that the number of agents required in the model to get an accurate estimation of mpox spread becomes quickly computationally prohibitive.

Finally, a major advantage of the network model of Van Djick et al. [8] is that it accounted for heterogeneity in contact patterns of transmission. Such an approach could be highly beneficial to study the spread of mpox in the context of the current outbreak as this would overcome the limitation of homogeneous mixing from compartmental models. Nevertheless, the applicability of network models in a broader context requires the use of recent contact data to adequately model the spread of the disease, and thus, there is a need for such studies of updated contact data that can adequately reflect the nature of current spread of the disease.

## Conclusion

This scoping review found that compartmental, branching process, Monte Carlo, agent-based, and network models have been utilized to study mpox transmission from a mathematical modelling perspective, with the first two model classes being the ones most commonly used. Each modelling strategy could be potentially useful to analyze aspects of the current outbreak depending of the level of complexity (e.g., compartmental models to analyze transmission over small regions, and stochastic or network models used for larger areas where variability could be larger).

However, the applicability of the models analyzed in this review to the current outbreak is currently limited by different underlying assumptions. The number and nature of compartments, homogenous mixing, the determination and usage of parameter values, the use of old and potentially obsolete or niche datasets, and the computational cost and complexity of the models are among the principal limitations of the different models to study the current mpox outbreak.

Overall, there is a need for new empirical epidemiological studies of mpox transmission that can provide data that is used to obtain estimates that are consistent in the context of the current outbreak. Additionally, all mathematical modelling strategies need to address the complexity in transmission due to the global mpox spread in non-endemic countries, where the nature of the interactions between environmental, social, and epidemiological factors is likely to have more ramifications than those existing in early mpox studies. Finally, the relatively low number of papers analyzed in this study reflects the very limited and sparse research in mpox, which highlights its historic condition as a neglected zoonosis.

The current outbreak has made clear that diseases which affect the global south need not be disregarded, as they can rapidly impact the rest of the world. More work needs to be done to ensure that in the future, resources and research are available to ensure the rapid management and containment of emerging zoonoses.

## Data Availability

All data produced in the present work are contained in the manuscript

## Credit authorship contribution statement

Jeta Molla: Conceptualization, Methodology, Identification and Screening of data, Software, Writing-original draft, Idriss Sekkak: Conceptualization, Identification and Screening of data, Methodology, Software, Writing-original draft., Ariel Mundo Ortiz: Screening data, Software, Writing-review & editing, Iain Moyles: Supervision, Writing-review & editing, Funding acquisition, Bouchra Nasri: Supervision, Software, Writing–review & editing, Funding acquisition.

## Acknowledgments

This work is supported by the Natural Sciences and Engineering Research Council of Canada (NSERC), RGPID-560523-2020, the NSERC OMNI EDIM, and NSERC Discovery grant no. 2019-06337

